# Patients with ACPA-positive and ACPA-negative Rheumatoid Arthritis Show Different Serological Autoantibody Repertoires and Autoantibody Associations with Disease Activity

**DOI:** 10.1101/2022.10.09.22280063

**Authors:** Kevin Y. Cunningham, Benjamin Hur, Vinod K. Gupta, Courtney A. Arment, Kerry A. Wright, Thomas G. Mason, Lynne S. Peterson, Delamo I. Bekele, Daniel E. Schaffer, Marissa L. Bailey, Kara E. Delger, Cynthia S. Crowson, Elena Myasoedova, Hu Zeng, Moses Rodriguez, Cornelia M. Weyand, John M. Davis, Jaeyun Sung

## Abstract

**Objectives:** Patients with rheumatoid arthritis (RA) can test either positive or negative for anti-citrullinated protein antibodies (ACPA), and are thereby ACPA-positive (ACPA+) or ACPA-negative (ACPA–), respectively. Through comprehensive profiling of autoantibodies in serum, we aimed to identify autoantibodies that are differentially abundant between patients with ACPA+ RA and ACPA– RA, and also those that are significantly associated with clinical disease activity.

**Methods:** Serum was collected from patients with ACPA+ RA (*n* = 32), ACPA– RA (*n* = 30), and healthy controls (*n* = 30). Sengenics Immunome^™^ protein microarray was used to screen for over 1,600 IgG autoantibodies against native, unmodified human proteins from each serum sample. Autoantibody profiles were compared between each RA subgroup and controls to identify differentially abundant autoantibodies (*P* < 0.05, Mann–Whitney *U* test; |Cliff’s delta (*d*)| > 0.33). Additionally, the relationship between RA patients’ autoantibody abundances and Clinical Disease Activity Index (CDAI) was examined for correlations between serum autoantibodies and disease activity (|Spearman’s *ρ*| > 0.4 and *P* < 0.01).

**Results:** We identified differences in serum autoantibodies between patients with ACPA+ RA and ACPA– RA compared with healthy controls. Specifically, we found 22 and 19 autoantibodies higher in ACPA+ RA patients and ACPA– RA patients, respectively. Among these two sets of autoantibodies, only one autoantibody (anti-GTF2A2) was common in both comparisons. On the other hand, we found 30 and 25 autoantibodies lower in ACPA+ RA and ACPA– RA, respectively, of which eight autoantibodies were common in both comparisons. Functional enrichment analysis of the protein antigens targeted by these autoantibodies showed an over-representation of a range of essential biological processes, including programmed cell death, metabolism, and signal transduction. Lastly, we found that autoantibodies correlate with CDAI, but associate differently depending on the presence or absence of ACPA.

**Conclusions:** ACPA status in patients with RA determines not only the composition of the serum autoantibody repertoire, but also the correlative relationships between autoantibodies and disease activity. Notably, many of the autoantibodies identified herein were reported for the first time. Our findings warrant further investigation into the immunological differences between these two RA subgroups, and shed new light on the possible need for different treatment approaches.

## 1. Introduction

Rheumatoid arthritis (RA) is characterized by chronic inflammation of the diarthrodial joints and extra-articular systems [1]. It is one of the most prevalent autoimmune disorders and affects ∼1% of the U.S. population [2]. RA is commonly diagnosed through a serological test for the presence of anti-citrullinated protein antibodies (ACPA), and patients testing positive for ACPA are collectively known as “ACPA-positive RA” (ACPA+ RA). However, Sieghart *et al*. found the presence of immunoglobulin G (IgG) ACPA in only ∼60% of RA patients [3]. Therefore, patients can test negative for ACPA yet still be clinically diagnosed with RA, and are thereby designated as “ACPA-negative RA” (ACPA– RA).

ACPA+ RA and ACPA– RA patients have been shown to display differences in their disease course [4–6] and response to treatment with disease-modifying anti-rheumatic drugs (DMARDs) [7]. Recent epidemiological evidence suggests that these two subgroups of RA are distinct subtypes with their own risk factors, etiologies, and treatment strategies [7–9].

Moreover, despite the stable incidence of RA over recent decades, the proportion of ACPA– RA cases has increased significantly [10]. Currently, the pathophysiological differences between the ACPA+ and ACPA– subgroups of RA are not well understood, thereby limiting the stratification of effective treatment strategies.

To identify biomolecular or cellular differences between ACPA+ RA and ACPA– RA, investigators have used high-dimensional and high-throughput molecular profiling approaches, such as metabolomics [11], single-cell RNA-seq on peripheral blood mononuclear cells and synovial tissue [12], flow cytometry immunophenotyping [13], and gut microbiome sequencing [14]. Complementing these multi-omics approaches, several recent studies have turned to serum autoantibody profiling to identify serological differences between the two RA subgroups. For example, in a study by Poulsen *et al*. using high-density protein microarray technology, the investigators identified 86 and 76 autoantibodies in the plasma of anti-cyclic citrullinated peptide (anti-CCP)+ RA and anti-CCP– RA patients, respectively, of which 61 were in common [15]. In another study by Poulsen *et al*., differences in plasma IgG antibody reactivity to a wide range of citrullinated human proteins were observed between the same subgroups of RA [16]. Both of their exploratory analyses, however, pooled together plasma samples of each study group prior to autoantibody profiling, and hence do not account for interindividual heterogeneity expected within groups. Finally, a study by Reed *et al*. examined the presence of ACPA, rheumatoid factor (RF), and anti-carbamylated-protein (anti-CarP) autoantibodies, and 36 other types of autoantibodies in anti-CCP2+ RA and anti-CCP2– RA patients [17]. The authors found that 43.6% of the seronegative (i.e., anti-CCP2 IgG−/IgM RF−) RA patients tested positive for one or a combination of ACPA, RF, and anti-CarP autoantibodies. That these patients tested positive for RA-associated autoantibodies suggests that a single autoantibody (e.g., ACPA) is not enough to explain the full clinical spectrum. Instead, RA, as well as its subgroups, are likely characterized by autoantibodies targeting a multitude of antigens, as shown by Li *et al*. [18].

Although ACPA is already part of the diagnostic criteria in RA, we posit that more disease-relevant, circulating autoantibodies can be discovered by profiling with much higher throughput. Certainly, with the concept of patient stratification (e.g., risks, clinical subsets) and precision medicine in mind, there is a clear need to detect autoantibodies comprehensively and simultaneously. In this study, using an autoantibody screening platform with high multiplexing capacity, we investigated serum autoantibody abundances in patients with ACPA+ RA and ACPA– RA, as well as in healthy controls.

## 2. Material and methods

### 2.1 Study participants, subject enrollment, and sample collection

The study population consisted of patients with RA attending the outpatient practice of the Division of Rheumatology at Mayo Clinic in Rochester, Minnesota. Eligibility required patients to be adults 18 years of age or older with a clinical diagnosis of RA by a rheumatologist based on the American College of Rheumatology/European League Against Rheumatism 2010 revised classification criteria for RA [19]. Patients were excluded if they did not comprehend English; were unable to provide written informed consent; or were members of a vulnerable population (e.g., incarcerated subjects). On the other hand, patients were eligible irrespective of the use of any particular medication and of disease duration. This led to a total of 62 patients fulfilling the eligibility criteria. Clinical and demographic data, including age, sex, race, body mass index (BMI), smoking status, the numbers of tender and swollen joints, patient and evaluator global assessments, C-reactive protein (CRP, mg/L), and results for rheumatoid factor (RF, IU/mL) and anti-CCP (U/mL), were collected from the electronic medical records.

Serum samples from RA patients were stored in our ongoing Mayo Clinic Rheumatology Biobank. This biorepository was created for long-term storage of diverse biological samples (e.g., serum, plasma, stool, white blood cells) from RA patients for use in research. In addition, serum samples from matched healthy donors attending the Mayo Clinic Biobank were used as controls. Subjects in the healthy control group were reported as not having any overt disease nor adverse symptoms at the time of sample collection. All three study groups (i.e., ACPA+ RA, ACPA– RA, and controls) were matched based on subjects’ age, sex, smoking status, race, and BMI (*P* < 0.05, Fisher’s exact test). Additionally, the two RA subgroups were matched by disease duration, RF status, and medication use (*P* < 0.05, Fisher’s exact test). All study participants provided written informed consent.

This study was approved by the Mayo Clinic Institutional Review Board (no. 14-000616 and no. 08-007049) in accordance with the Declaration of Helsinki. All methods and procedures were performed in accordance with the Mayo Clinic Institutional Review Board guidelines and regulations. This manuscript follows the STrengthening the Reporting of OBservational studies in Epidemiology (STROBE) reporting guidelines for observational studies [20].

### 2.2 Clinical Disease Activity Index

The Clinical Disease Activity Index (CDAI) is a numeric scale used to measure disease activity in RA. CDAI is calculated by the summation of four individual elements: Swollen Joint Count (0–28), Tender Joint Count (0–28), Patient Global Assessment of Disease Activity (0–10.0), and Evaluator Global Assessment of Disease Activity (0–10.0) [21]. The full range of CDAI is 0–76. CDAI can be used to categorize RA disease activity into four states: Remission (CDAI ≤ 2.8), Low Disease Activity (2.8 < CDAI ≤ 10), Moderate Disease Activity (10 < CDAI ≤ 22), and High Disease Activity (CDAI > 22).

### 2.3 Autoantibody microarray protocol

#### 2.3.1 Serum dilution

IgG autoantibody profiling of all serum samples was performed by Sengenics (Singapore) Immunome^™^ Protein Microarray. The microarray is comprised of quadruplicate spots of 1,622 full-length, correctly folded, and native (i.e., not post-translationally modified) human proteins, which are immobilized through a proprietary biotin carboxyl carrier protein (BCCP) folding marker. These proteins cover various protein families, including kinases, signaling molecules, cytokines, and transcription factors. Frozen serum samples were randomized before being assigned to assay racks. Samples were stored at −20°C during experimental setup. Each sample was thawed in a shaking incubator at +20°C for 30 minutes, vortexed three times at full speed, and then spun down for three minutes at 13,000 RPM in a microcentrifuge. Next, 5.625 μL of the sample was pipetted into 4.5 mL of Serum Assay Buffer (SAB). The buffer contained 0.1% v/v Triton, 0.1% w/v BSA in 1X phosphate-buffered saline (PBS) (20°C) and vortexed three times. The tube was tilted during aspiration to ensure that the serum was sampled below the lipid layer at the top but not from the bottom of the tube in case of any sediment. This serum dilution process was carried out in a class II biological safety cabinet. Batch records were marked accordingly to ensure that the correct samples were added to the tubes.

#### 2.3.2 Biomarker assay

The array was removed from the storage buffer, placed in a slide box and rack with 200 mL cold SAB, and shaken on an orbital shaker at 50 RPM for five minutes. After the slides were rinsed entirely, they were placed with the array side up in a slide hybridization chamber with individual sera that had been diluted earlier. All slides were scanned and incubated on a horizontal shaker at 50 RPM for two hours at 20°C.

#### 2.3.3 Array washing after serum binding

The protein array slide was then rinsed twice in individual “Pap jars” with 30 mL SAB, followed by 200 mL of SAB buffer in the slide staining box for 20 minutes on the shaker at 50 RPM at room temperature. All slides were transferred sequentially and in the same orientation.

#### 2.3.4 Incubation with Cy3-anti IgG

Binding of IgG was detected by incubation with Cy3-rabbit anti-human IgG (Dako Cytomation) labeled according to the manufacturer’s recommended protocols (GE Healthcare). Arrays were then immersed in a hybridization solution containing a mixture of Cy3-rabbit anti-human IgG solution diluted 1:1000 in SAB buffer for 2 hours at 50 RPM in 20°C.

#### 2.3.5 Washing after incubation with Cy3-anti IgG

After the incubation, each slide was dipped in 200 mL of SAB buffer three times for five minutes at 50 RPM at room temperature. Excess buffer was removed by immersing the slide in 200 mL of pure water for a few minutes. Slides were dried for four minutes and stored at room temperature until scanning on the same day. Hybridization signals were measured with a microarray laser scanner (Agilent Scanner) at 10 μm resolution. Fluorescence levels were detected according to the manufacturer’s instructions, whereby each spot is plotted using Agilent Feature Extraction software.

### 2.4 Image analysis and data extraction

To extract quantitative data from the slides, image analysis was utilized to evaluate the number of autoantibodies present in each serum sample by measuring the median intensities of all the pixels within each probed spot. A raw .tiff image file was generated for each slide (sample). Automatic extraction and quantification of all the pixels in each spot on the array were performed using GenePix Pro 7 software (Molecular Devices), which provides statistics for each probed spot on the array. This includes the mean and median of the pixel intensities within a spot and its local background. A GAL (GenePix Array List) file for the array was generated to aid the image analysis. This file contains the information of all probed spots and their positions on the array. Following data extraction, a GenePix Results (.GPR) file, which contains information for each spot (e.g., Protein ID, protein name, foreground intensities, background intensities), was generated for each slide.

### 2.5 Data handling and pre-processing

The quadruplicate spots for each antibody were measured and averaged for each slide (**Supplementary Information** and **Supplementary Fig. 1**). This resulted in a data sheet that contains both foreground and background intensities of each spot represented in relative fluorescence units (RFUs). Raw data can be found in **Supplementary Table 1**. Raw RFU values from the microarrays were quantile normalized before all analyses described below.

### 2.6 Feature selection prior to principal component analysis

Principal component analysis (PCA) was used to project the autoantibody profiles onto an ordination plot. Autoantibody features deemed invariant across all study participants were removed prior to PCA, as they were assumed to not significantly contribute to the underlying variance in the dataset. For this, a one-way ANOVA test was performed on each autoantibody feature across all profiles, and the subset of those that were statistically significant (*P* < 0.05) was utilized for PCA.

### 2.7 Identifying differentially abundant autoantibodies

An autoantibody was considered differentially abundant between two study groups when found to be statistically significant (*P* < 0.05, Mann–Whitney *U* test) with medium effect size (|Cliff’s delta (*d*)| > 0.33, as defined in [22]). Cliff’s delta, a non-parametric measure of effect size, tells how often values in one group are larger than the values in the second group.

### 2.8 Functional enrichment analysis of autoantigens

For a set of autoantibodies, functional enrichment analysis was performed on the Gene Ontology Biological Process (GOTERM_BP_FAT) annotations of their target antigens using the DAVID online tool [23]. A *P*-value of 0.05 from a modified one-tailed Fisher’s exact test was used as the significance cutoff.

### 2.9 Spearman correlations between Clinical Disease Activity Index and autoantibody abundances

The Spearman correlation coefficient *ρ* was used to measure the strength of the relationship between the patient CDAI scores and autoantibody abundances. |*ρ*| > 0.4 and *P*-value < 0.01 were chosen as the significance cutoffs.

### 2.10 Data availability

Source code and data sets used to generate the results presented in this study are available at: https://github.com/jaeyunsung/RA_Autoantibodies_2022.

## 3. Results

### 3.1 Study population and clinical/demographic characteristics

An overview of our study design is presented in **Figure 1A** and is described as follows: This retrospective, observational cohort study includes a total of 92 participants comprised of three study groups, i.e., patients with ACPA+ RA (*n* = 32), patients with ACPA– RA (*n* = 30), and healthy controls (*n* = 30). At the time of serum sample collection, all RA patients had established disease with a mean age of 62.2 years (range: 45–75 years); a mean disease duration of 8.1 years (range: 1–26 years); 69.4% (43 of 62) were female; and disease activity of patients varied from remission to high disease activity, with a mean Clinical Disease Activity Index (CDAI) of 10.6 (range: 0–66.8). Subsets of patients were on treatment with biologic disease-modifying anti-rheumatic drugs (bDMARDs) (33.9% or 21 of 62), conventional synthetic disease-modifying anti-rheumatic drugs (csDMARDs) (82.3% or 51 of 62), targeted synthetic disease-modifying anti-rheumatic drugs (tsDMARDs) (4.8% or 3 of 62), or prednisone (33.9% or 21 of 62). **Table 1** and **Supplementary Tables 2** and **3** provide the clinical and demographic characteristics of the study groups and study participants, respectively. Serum samples collected from all participants underwent comprehensive profiling of 1,622 IgG autoantibodies using the Sengenics Immunome^™^ Protein Microarray technology.

**Figure 1.**
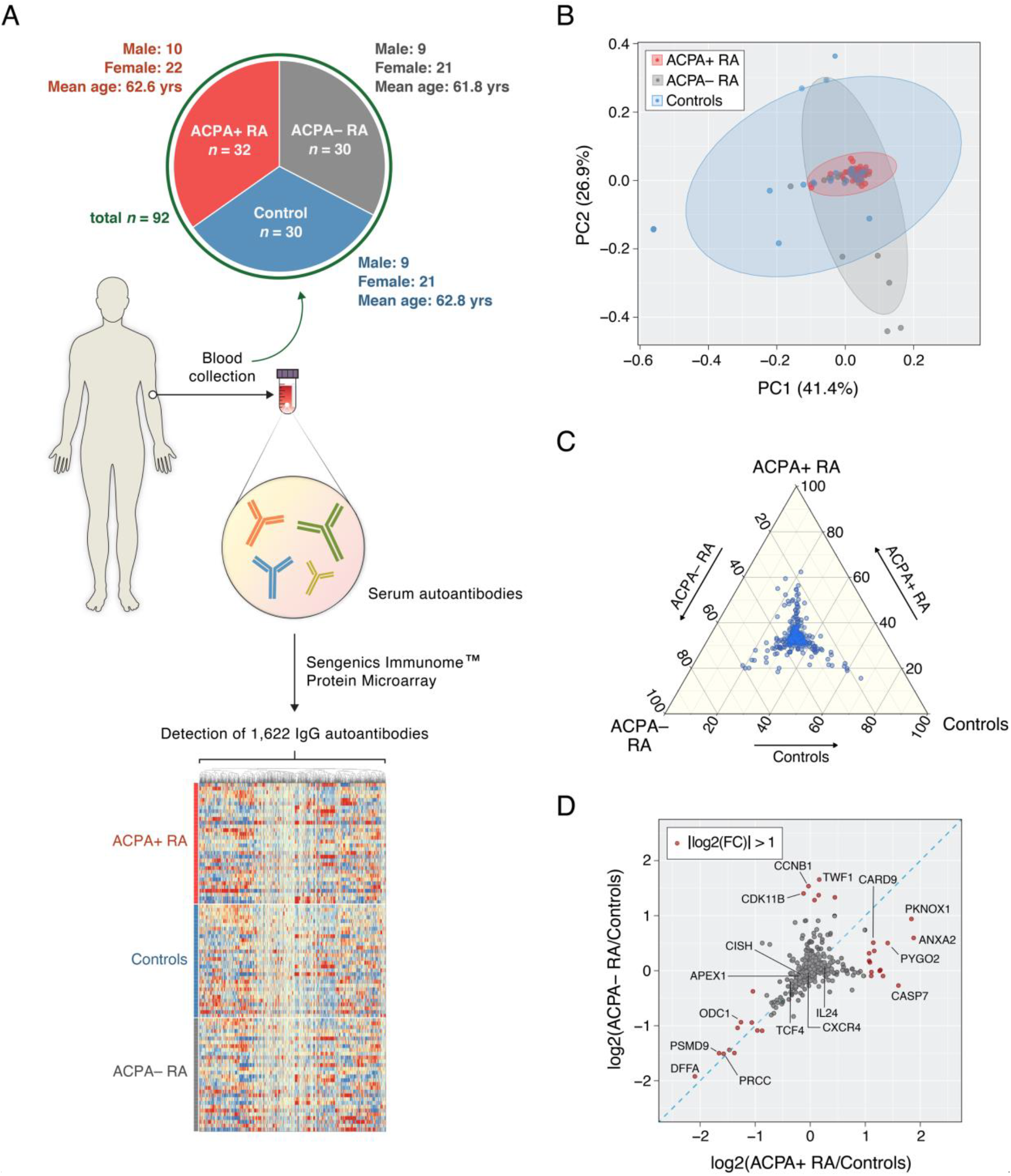
Group-wise comparisons of serum autoantibody composition profiles. (**A**) Blood (serum) samples were collected to examine autoantibody compositions in ACPA+ RA (*n* = 32), ACPA– RA (*n* = 30), and healthy controls (*n* = 30). By using the Sengenics Immunome^™^ Protein Microarray, each serum sample was screened for 1,622 IgG autoantibodies that target human immune-related proteins in their native conformations. The heatmap illustrates autoantibodies clustered according to abundance similarities across samples. (**B**) Ordination plot of principal component analysis on the autoantibody profiles. (**C**) Ternary plot showing normalized mean abundances of 1,622 autoantibodies across ACPA+ RA, ACPA– RA, and controls. Coordinates of each point correspond to percentages of mean abundances and sum to 100. (**D**) Fold-changes in mean autoantibody abundances between an RA subgroup and the control group. X-axis and y-axis correspond to the fold-changes between ACPA+ RA and controls and between ACPA– RA and controls, respectively. Points shown in red represent autoantibodies that have a fold-change of 2 (or greater) between an RA subgroup and controls. The blue diagonal dashed line represents the line y=x. For brevity, the points in the scatterplot are labeled by the names of the autoantibody targets.

**Table 1.**
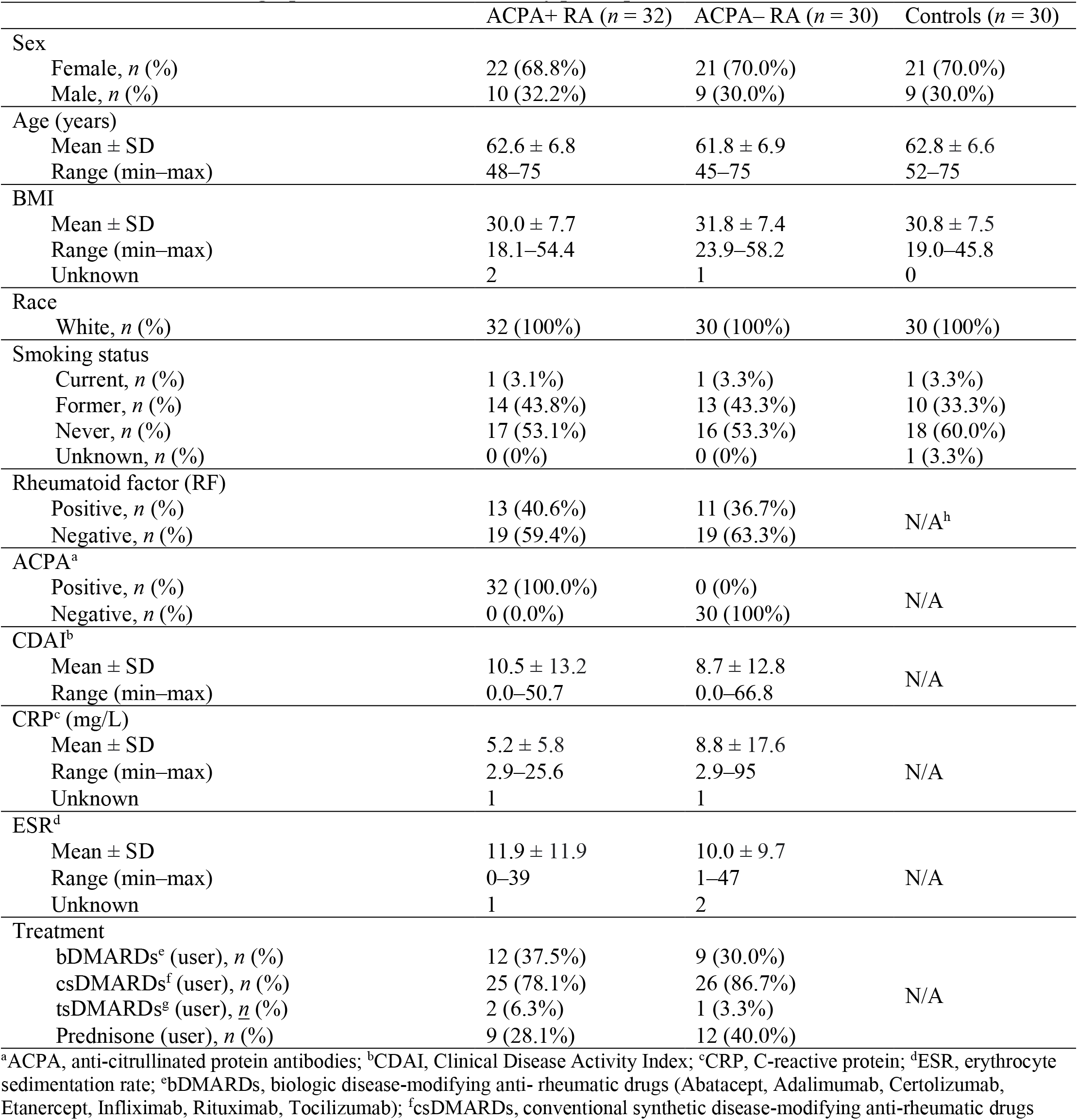
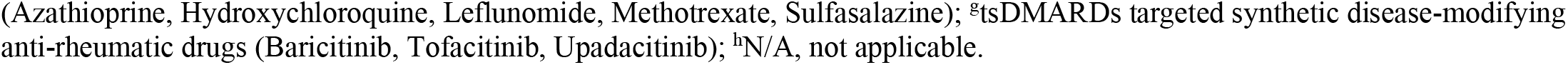
Clinical and demographic characteristics of study participants.

|  | ACPA+ RA (*n* = 32) | ACPA– RA (*n* = 30) | Controls (*n* = 30) |
| --- | --- | --- | --- |
| Sex |  |  |  |
| Female, *n* (%) | 22 (68.8%) | 21 (70.0%) | 21 (70.0%) |
| Male, *n* (%) | 10 (32.2%) | 9 (30.0%) | 9 (30.0%) |
| Age (years) |  |  |  |
| Mean ± SD | 62.6 ± 6.8 | 61.8 ± 6.9 | 62.8 ± 6.6 |
| Range (min–max) | 48–75 | 45–75 | 52–75 |
| BMI |  |  |  |
| Mean ± SD | 30.0 ± 7.7 | 31.8 ± 7.4 | 30.8 ± 7.5 |
| Range (min–max) | 18.1–54.4 | 23.9–58.2 | 19.0–45.8 |
| Unknown | 2 | 1 | 0 |
| Race |  |  |  |
| White, *n* (%) | 32 (100%) | 30 (100%) | 30 (100%) |
| Smoking status |  |  |  |
| Current, *n* (%) | 1 (3.1%) | 1 (3.3%) | 1 (3.3%) |
| Former, *n* (%) | 14 (43.8%) | 13 (43.3%) | 10 (33.3%) |
| Never, *n* (%) | 17 (53.1%) | 16 (53.3%) | 18 (60.0%) |
| Unknown, *n* (%) | 0 (0%) | 0 (0%) | 1 (3.3%) |
| Rheumatoid factor (RF) |  |  |  |
| Positive, *n* (%) | 13 (40.6%) | 11 (36.7%) | N/A<sup>h</sup> |
| Negative, *n* (%) | 19 (59.4%) | 19 (63.3%) |  |
| ACPA<sup>a</sup> |  |  |  |
| Positive, *n* (%) | 32 (100.0%) | 0 (0%) | N/A |
| Negative, *n* (%) | 0 (0.0%) | 30 (100%) |  |
| CDAI<sup>b</sup> |  |  |  |
| Mean ± SD | 10.5 ± 13.2 | 8.7 ± 12.8 | N/A |
| Range (min–max) | 0.0–50.7 | 0.0–66.8 |  |
| CRP<sup>c</sup> (mg/L) |  |  |  |
| Mean ± SD | 5.2 ± 5.8 | 8.8 ± 17.6 | N/A |
| Range (min–max) | 2.9–25.6 | 2.9–95 |  |
| Unknown | 1 | 1 |  |
| ESR<sup>d</sup> |  |  |  |
| Mean ± SD | 11.9 ± 11.9 | 10.0 ± 9.7 | N/A |
| Range (min–max) | 0–39 | 1–47 |  |
| Unknown | 1 | 2 |  |
| Treatment |  |  |  |
| bDMARDs<sup>e</sup> (user), *n* (%) | 12 (37.5%) | 9 (30.0%) | N/A |
| csDMARDs<sup>f</sup> (user), *n* (%) | 25 (78.1%) | 26 (86.7%) |  |
| tsDMARDs<sup>g</sup> (user), *n* (%) | 2 (6.3%) | 1 (3.3%) |  |
| Prednisone (user), *n* (%) | 9 (28.1%) | 12 (40.0%) |  |

<sup>a</sup>ACPA, anti-citrullinated protein antibodies; <sup>b</sup>CDAI, Clinical Disease Activity Index; <sup>c</sup>CRP, C-reactive protein; <sup>d</sup>ESR, erythrocyte sedimentation rate; <sup>e</sup>bDMARDs, biologic disease-modifying anti-rheumatic drugs (Abatacept, Adalimumab, Certolizumab, Etanercept, Infliximab, Rituximab, Tocilizumab); <sup>f</sup>csDMARDs, conventional synthetic disease-modifying anti-rheumatic drugs
(Azathioprine, Hydroxychloroquine, Leflunomide, Methotrexate, Sulfasalazine); <sup>g</sup>tsDMARDs targeted synthetic disease-modifying anti-rheumatic drugs (Baricitinib, Tofacitinib, Upadacitinib); <sup>h</sup>N/A, not applicable.

### 3.2 Serum autoantibody profiles in ACPA+ RA, ACPA– RA, and healthy controls

Our PCA results show that all three study groups display within (intra)-group heterogeneity in their serum autoantibody profiles (**Fig. 1B**). Controls showed the largest heterogeneity in autoantibody composition. In contrast, the ACPA+ RA group showed the smallest heterogeneity, possibly indicating that ACPA+ RA is a more uniform disease subgroup based on immunological features. In a ternary plot showing the normalized mean abundances of individual autoantibodies across the three study groups (**Fig. 1C** and **Supplementary Table 4**), we observed that most autoantibodies had similar mean abundances. However, several autoantibodies were found to have noticeably higher abundances in a particular study group (see points closer to the corners), possibly representing group-specific characteristics. Finally, when examining the autoantibody abundances in ACPA+ RA and ACPA– RA in relation to controls (**Supplementary Table 5**), we again observed autoantibodies whose abundances uniquely characterize a specific study group (**Fig. 1D**, see points in red). In all, our results suggest that not only do patients with ACPA+ RA and ACPA– RA portray serological differences in autoantibody abundances, but also lower abundances of certain autoantibodies (compared with controls) could potentially be a novel hallmark of RA.

### 3.3 Identifying differentially abundant autoantibodies in RA subgroups

We next aimed to characterize the differences in serum autoantibody abundances between ACPA+ RA and ACPA– RA in further detail. We identified 22 and 19 autoantibodies significantly higher in ACPA+ RA and ACPA– RA, respectively, compared with controls (**Fig. 2** and **Supplementary Table 6**). A notable target of an autoantibody higher in ACPA+ RA is IL24. In a study involving patients with RA, osteoarthritis, and spondyloarthropathy, Kragstrup *et al*. found higher levels of IL24 in synovial fluid and plasma of RA and spondyloarthropathy patients compared with osteoarthritis patients [24]. In light of our findings, higher abundances of anti-IL24 autoantibodies in the ACPA+ subgroup of RA could indicate a possible route for the body to compensate for an overabundance of IL24. The only autoantibody found to be higher in both RA subgroups compared with controls was for GTF2A2. Although the role of GTF2A2 in RA is currently unknown, it has been previously found in systemic lupus erythematosus (SLE) that point mutations in the expression quantitative trait loci (eQTL) of this transcription factor subunit are associated with type I interferon levels [25].

**Figure 2.**
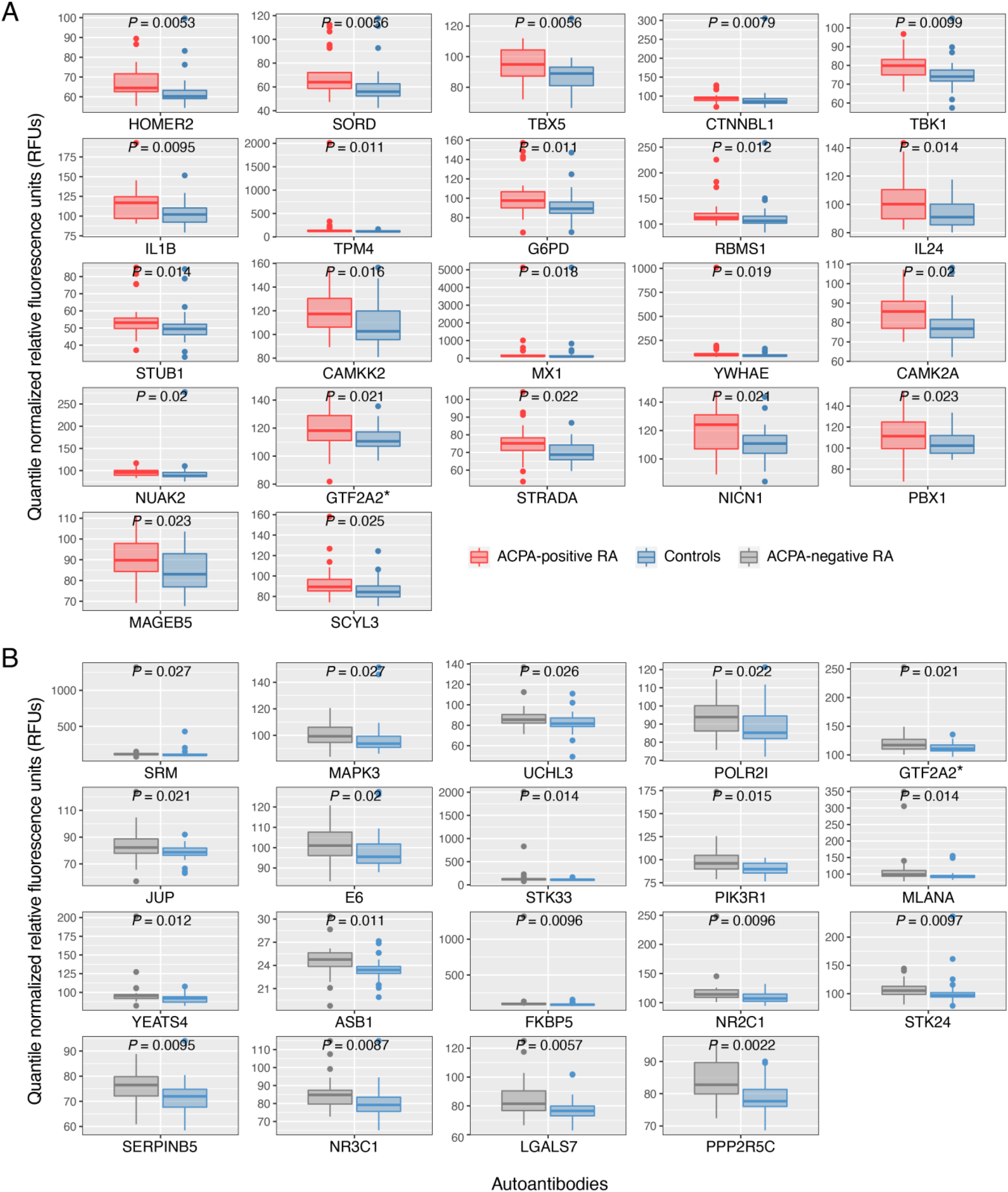
Serum autoantibodies found to have significantly higher abundances in ACPA+ RA and ACPA– RA compared with healthy controls. Patients with **(A)** ACPA+ RA and **(B)** ACPA– RA show higher abundances in 22 and 19 autoantibodies, respectively. Two-sided Mann–Whitney *U* test (*P* < 0.05) and the Cliff’s Delta effect size (|*d*| > 0.33) were used to find autoantibodies of significantly higher abundance. Standard box-and-whisker plots (e.g., center line, median; box limits, upper and lower quartiles; whiskers, 1.5× interquartile range; points, outliers) are used to show autoantibody abundances. *, autoantibody for GTF2A2 was found to have significantly higher abundances in both ACPA+ RA and ACPA– RA subgroups. For brevity, the plots are labeled by the names of the autoantibody targets.

In contrast to the autoantibodies higher in RA, we next sought to identify autoantibodies that were lower in either RA subgroup. In other words, could there be circulating autoantibodies that were higher in healthy and presumably integral to normal physiology and immune homeostasis? For the first time, we report 30 and 25 autoantibodies that were significantly lower in ACPA+ RA and ACPA– RA, respectively, compared with controls (**Fig. 3** and **Supplementary Table 7**). Among these two sets of identified autoantibodies, eight were in common and target APEX1, DAPK2, MAP4, PSMD4, SIK2, SOCS5, STAM2, and TCF4. Interestingly, the transcription factor TCF4 has been suggested as a potential therapeutic target for osteoarthritis [26].

**Figure 3.**
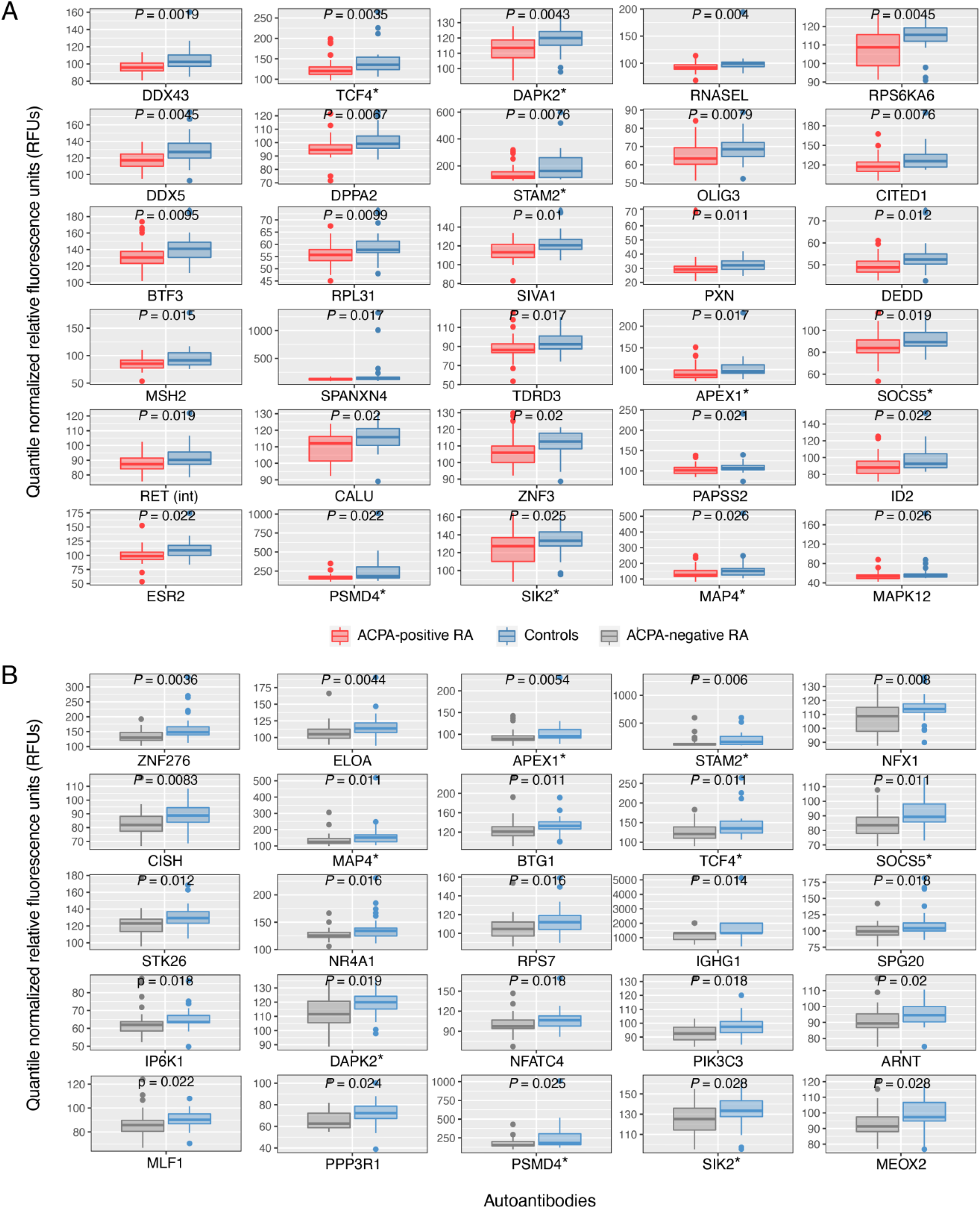
Serum autoantibodies found to have significantly lower abundances in ACPA+ RA and ACPA– RA compared with healthy controls. Patients with **(A)** ACPA+ RA and **(B)** ACPA– RA show lower abundances in 30 and 25 autoantibodies, respectively. Two-sided Mann–Whitney *U* test (*P* < 0.05) and Cliff’s Delta effect size (|*d*| > 0.33) were used to find autoantibodies of significantly lower abundance. Standard box-and-whisker plots (e.g., center line, median; box limits, upper and lower quartiles; whiskers, 1.5× interquartile range; points, outliers) are used to show autoantibody abundances. *, autoantibodies common to both ACPA+ RA and ACPA– RA subgroups. For brevity, the box-and-whisker plots are labeled by the names of the autoantibody targets.

### 3.4 Functional associations of the autoantibody targets

Having identified differentially abundant autoantibodies in both subgroups of RA, we next set out to characterize the functions of the targeted autoantigens. Functional enrichment (GO terms) using DAVID (see subsection 2.8 in **Material and methods**) found that the top enriched biological processes of the targets covered a range of fundamental cellular functions, including programmed cell death, transcription, metabolism and biosynthesis, and signal transduction (**Fig. 4**). We identified 55 and 38 enriched biological processes from the targets of differentially abundant autoantibodies higher in ACPA+ RA and ACPA– RA, respectively (*P* < 0.05, **Fig. 4A** and **Supplementary Table 8**). The top three enriched biological processes of the targets of autoantibodies higher in ACPA+ RA were Cell Death, Apoptotic Process, and Programmed Cell Death (**Fig. 4A**, blue bars), possibly implicating dysregulated programmed cell death in ACPA+ RA. Apoptosis, a normal process of programmed cell death, is critical in regulating and maintaining tissue growth and homeostasis [27]. Although apoptosis is not known to induce an inflammatory response in the absence of disease, disruptions in apoptotic pathways have been reported in autoimmunity [28], such as in SLE [29]. For the targets of autoantibodies higher in ACPA– RA, we found that the most highly enriched biological processes were related to gene expression, including DNA-templated Transcription, Transcription Initiation from RNA Polymerase II Promoter, and Cellular Response to Endogenous Stimulus (**Fig. 4A**, orange bars).

**Figure 4.**
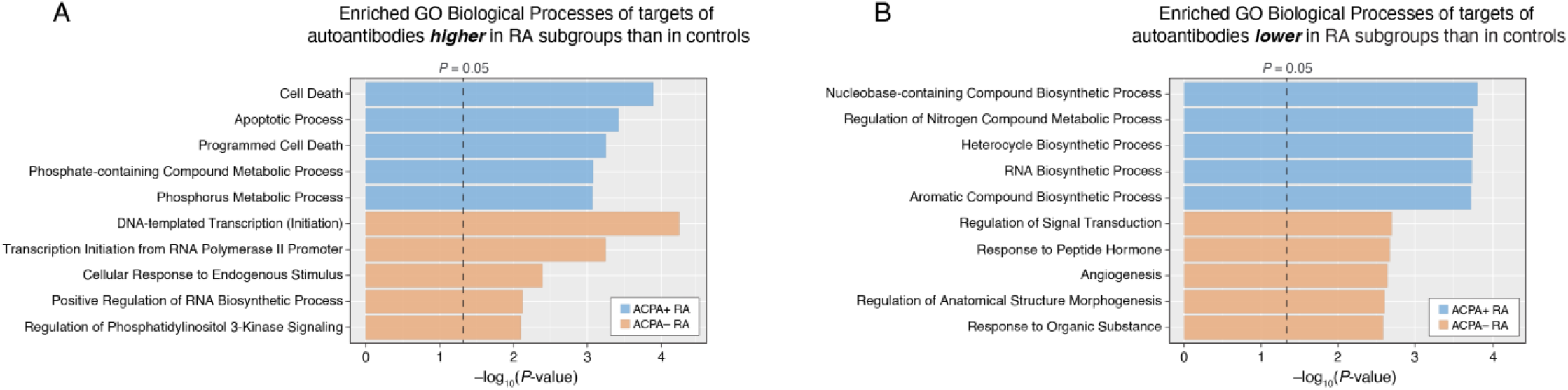
Autoantigens targeted by the differentially abundant autoantibodies show enrichment in fundamental cellular functions. The top 5 statistically enriched biological processes of targets of autoantibodies that were significantly **(A)** higher and **(B)** lower in RA subgroups (ACPA+, ACPA–) compared with healthy controls. Enriched (over-represented) biological processes were rank-ordered based on the modified one-tailed Fisher’s exact test *P*-values provided in DAVID.

We identified 76 and 92 enriched biological processes from the targets of differentially abundant autoantibodies lower in ACPA+ RA and ACPA– RA, respectively (*P* < 0.05, **Fig. 4B** and **Supplementary Table 9**). Notably, all of the top 5 enriched biological processes for targets whose autoantibodies were lower in ACPA+ RA were related to either biosynthetic or metabolic processes: Nucleobase-containing Compound Biosynthetic Process, Regulation of Nitrogen Compound Metabolic Process, Heterocycle Biosynthetic Process, RNA Biosynthetic Process, and Aromatic Compound Biosynthetic Process (**Fig. 4B**, blue bars). Additionally, the top 5 enriched biological processes for targets whose autoantibodies were lower in ACPA– RA were related to cell signaling (Regulation of Signal Transduction), cellular response (Response to Peptide Hormone, Angiogenesis), and tissue structure (Regulation of Anatomical Structure Morphogenesis) (**Fig. 4B**, orange bars). Interestingly, regarding signal regulation and cellular response, an imbalance of G-protein-coupled receptor (GPCR)-specific autoantibody levels was found to be associated with autoimmune disorders, including RA and SLE [30]. Unraveling how autoantibodies react to, and thereby dysregulate, these essential cellular functions in RA may provide new avenues for identifying tractable therapeutic targets.

### 3.5 Autoantibody abundances are significantly correlated with RA disease activity

To determine whether serum autoantibodies reflect disease activity in RA, we examined for autoantibodies correlated with the CDAI (see subsection 2.9 in **Material and methods**). To the best of our knowledge, we report for the first time 27 autoantibodies that have at least a moderate correlation with CDAI (|Spearman’s *ρ*| > 0.4 and *P* < 0.01) in either or both subgroups of RA (**Fig. 5** and **Supplementary Table 10**). Furthermore, we found that these correlations differed based on ACPA status, providing further evidence of serological differences between the two RA subgroups. Specifically, eleven of the 27 autoantibodies were significantly correlated with CDAI in ACPA+ RA, with six (for PYGB, EXT2, CDKN2B, FAS, GNA15, and MMP2) being positively correlated, and five (for MED4, RAB38, PAK2, AK1, and PELO) being negatively correlated. In addition, fifteen among the 27 were significantly correlated with CDAI in ACPA– RA, with eight (for SP1, TPM3, FRK, ELK1, CLK3, TPM1, DDIT3, and MARK3) and seven (for VDR, CAPG, AHSG, CXCR4, EGR2, DCLK1, and ESR2) having positive and negative correlations with CDAI, respectively. Finally, three (for PELO, CLK3, and CISH) of the 27 autoantibody abundances were significantly correlated with CDAI in all RA patients, with a subset of those already found to be significant in ACPA+ RA (for PELO) and ACPA– RA (for CLK3). Autoantibodies for Cytokine inducible SH2 containing protein (CISH) were positively correlated with CDAI (*ρ* = 0.44, *P* = 6.5 × 10^×4^) when considering both subgroups simultaneously (**Supplementary Fig. 2**), but not when taking into account the individual subgroups of RA.

**Figure 5.**
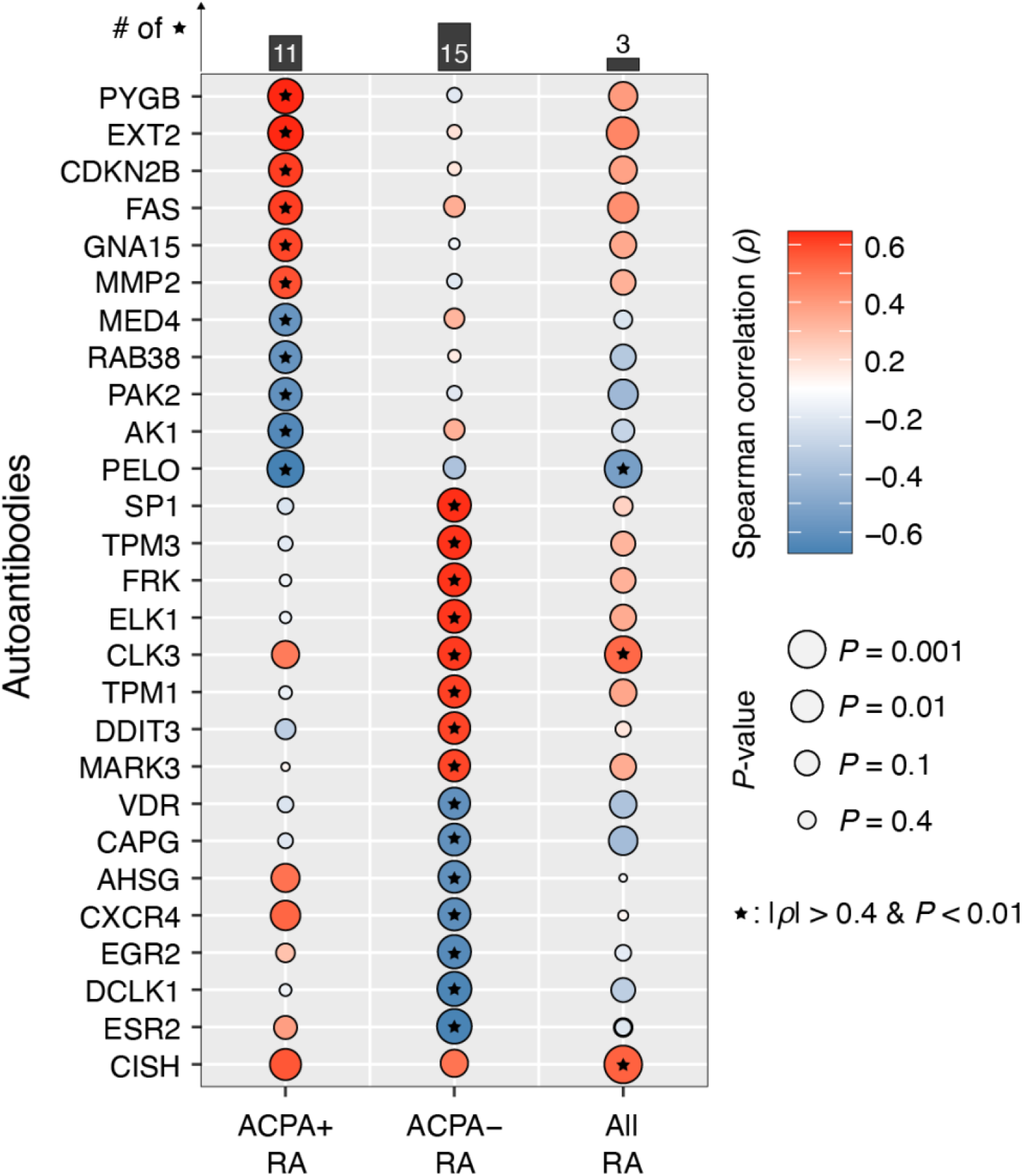
Serum autoantibodies in RA patients display significant correlations with CDAI. The strength of the relationships between autoantibody abundances and CDAI was measured in three groups: ACPA+ RA, ACPA– RA, and all RA patients. 27 different autoantibodies were significantly correlated with CDAI (|Spearman’s *ρ*| > 0.4 and *P* < 0.01). For brevity, the autoantibodies are labeled by the name of their targets.

The connections between serum autoantibodies and RA disease activity mentioned above have not been reported in prior studies. However, several of their protein targets (autoantigens) have been previously linked to RA. For example, we identified autoantibody abundances for Fas cell surface death receptor (FAS) as being positively correlated with CDAI in ACPA+ RA patients (*ρ =* 0.511, *P* = 0.005). The interaction of this receptor with its ligand is known to initiate a signaling cascade that leads to apoptosis [31]. Chou *et al*. found that FAS proteins in synovial tissue were higher in patients with RA than in patients with osteoarthritis and post-traumatic joint disease [32]. In addition, we found that autoantibodies for CXC chemokine 4 (CXCR4) displayed a negative correlation with CDAI in ACPA– RA patients (*ρ =* -0.508, *P* = 0.007). Peng *et al*. found that CXCR4 levels in serum and joint synovial fluid strongly correlated with RA patients’ disease activity score-28 (DAS28) [33]. Our findings associating RA disease activity with anti-FAS and anti-CXCR4 autoantibodies support previous reports regarding the relevance of FAS and CXCR4 in RA.

## 4. Discussion

This study addresses the following key questions: Are there differences in serum autoantibody abundances between ACPA+ RA and ACPA– RA? In particular, which serum autoantibodies in the two RA subgroups are either higher or lower in abundance compared with healthy controls? Which biological functions are the autoantibody targets involved in? Do any of the circulating autoantibodies correlate with disease activity in RA patients? To these ends, we used a multiplex autoantibody screening array on the serum of patients with ACPA+ RA and ACPA– RA. Of note, multiplex autoantibody profiling has newly identified autoantibodies associated with SLE [34,35], COVID-19 [36], and cancers [37–39]. Our statistical analysis of these data identified differentially abundant autoantibodies between subgroups of RA and healthy controls. Interestingly, anti-GTF2A2 was the only autoantibody found to be higher in both ACPA+ RA and ACPA– RA; this provides further evidence of previously reported immunological differences between these two subgroups despite sharing similar symptoms [8,40], although the precise pathophysiological mechanisms remain yet unclear. Functional enrichment analysis on the autoantibody targets showed that the autoantibodies could be involved in—and possibly interfering with—various essential biological processes, and differently so depending on the RA subgroup. Finally, not only did we find that serum autoantibodies correlate with RA disease activity, but also we report for the first time that these correlations differ based on ACPA status.

It has long been established that the presence or elevation of autoantibodies in circulation is a hallmark of RA [41]. Despite this commonly known attribute of RA, we found a total of 47 unique autoantibodies lower in both RA subgroups compared with controls, possibly suggesting “missing” features of health; however, the reason for this finding is not known at this point. Notably, there were eight common autoantibodies lower in both ACPA+ RA and ACPA– RA, including anti-APEX1, anti-STAM2, and anti-SOCS5. These could be natural autoantibodies (which are produced by the immune system to intentionally target self-antigens) that aid in regulating normal homeostatic processes [42], clearing cellular debris and other waste products from dying cells [43], catalyzing enzymatic reactions [44], and training the adaptive immune system to protect against self-antigens that may induce a severe immune response [45]. Notably, the protective function of natural autoantibodies has been generally ascribed to the IgM class [46]; nevertheless, it is now beginning to be discovered that an IgG autoantibody repertoire exists in the blood of healthy individuals [47,48] that is considered to be highly individualized and stable in adults [47]. Examining whether the IgG autoantibodies lower in RA subgroups play any causal role in attenuating RA severity, or whether any of these may be utilized for therapeutic purposes, would be appealing topics for future studies.

As we demonstrate in our study, comprehensive serological profiling of autoantibodies can be instrumental in providing novel insights into potentially pathogenic or disease-modulating autoantibodies (or the autoantigens) in RA; and into how the disruption of natural autoantibody production might be connected to the disease. However, several limitations should be acknowledged when interpreting our results. First, the relatively small sample sizes are likely not enough to fully represent the intricate characteristics of each study group, so future works will need to include considerably more participants. Second, and in a related vein, our study is not a broad representation of patients with RA, as all study participants were Caucasian mostly from the Midwest region of the United States. Thus we emphasize caution when extending our findings to patients of different racial backgrounds, considering disparities in RA prevalence found among different races [49]. Third, despite the multi-analyte detection strategy we took to simultaneously profile >1,600 autoantibodies, our entire results are still based on measurements from a single technique. Therefore, our findings may not be fully replicable across different autoantibody profiling platforms. Especially for diagnostic biomarker discovery purposes, our results will require validation using traditional methods such as ELISA and western blotting. Fourth, the microarray used in this study provides quantitative estimates of autoantibodies that recognize human proteins in only their native, unmodified state. Performing a similar analysis with autoantibodies reactive against post-translationally modified (e.g., (homo)citrullination, carbamylation) proteins would be an intriguing future direction. Fifth, we lose most of the significant (*P* < 0.05, Mann–Whitney *U*) “hits” after Benjamini–Hochberg correction. This could be attributed to multiple factors including a lack of strong differences (in serum autoantibody abundances) between study groups and the large number of tests resulting from the high dimensionality of our screening platform. Despite their modest statistical significance, the observed differences nevertheless passed an additional criterion concerning the effect size (|*d*| *>* 0.33).

Finally, given the observational nature of this study, we cannot establish at this point whether the identified autoantibodies (or autoantigens) are actually involved in inflammation or other symptoms of RA. Deciphering how the autoantibodies are linked to pathogenic events at the site of disease (synovial fluid) lies at the forefront of our future research.

The autoantibodies reported herein are expected to motivate future studies examining their potential as useful clinical biomarkers in RA, especially as we move toward classifying patients based on their molecular features [50,51]. In this regard, machine learning can translate large-scale datasets into more actionable information for RA, e.g., risk factors or predictors of disease course, as demonstrated in our previous works [52–54]. Of note, there is yet no diagnostic laboratory test to confirm RA and differentiate it from other inflammatory arthritides in the absence of ACPA; in effect, ACPA– RA patients tend to be diagnosed well after the onset of disease, leading to delays in starting treatment and suboptimal long-term clinical outcomes [55]. Identifying blood biomarkers specifically in ACPA– RA could greatly benefit patients by contributing to an earlier diagnosis, as a timely and focused patient management plan can limit disease progression and preserve the quality of life [56].

## Data Availability

Source code and data sets used to generate the results presented in this study are available at:
https://github.com/jaeyunsung/RA_Autoantibodies_2022.

## Acknowledgments

We thank all study participants who volunteered for this study, especially our dear patients. We also thank the Mayo Clinic Division of Rheumatology study coordinators and the Mayo Clinic Biobank staff for facilitating serum sample identification, packaging, and shipment. This work was supported by a gift from Mark E. and Mary A. Davis to the Mayo Clinic Center for Individualized Medicine. The funding source of this study was not involved in the study design; in the collection, analysis, and interpretation of the data; in the writing of the manuscript; and in the decision to submit the article for publication.

## Author Contributions

All authors were involved in drafting and revising the article, or contributing important intellectual content. All authors approved the final version to be published. Dr. Sung has full access to all data in the study and takes responsibility for the integrity and accuracy of the data analysis.

Kevin Y. Cunningham: Data curation, Formal analysis, Investigation, Software, Visualization, Writing - original draft, Writing - review & editing.

Benjamin Hur: Software, Formal analysis, Writing - review & editing

Vinod K. Gupta: Formal analysis, Writing - review & editing

Courtney A. Arment: Resources

Kerry A. Wright: Resources

Thomas G. Mason: Resources

Lynne S. Peterson: Resources

Delamo I. Bekele: Resources

Daniel E. Schaffer: Resources

Marissa L. Bailey: Resources

Kara E. Delger: Resources

Cynthia S. Crowson: Writing - review & editing

Elena Myasoedova: Writing - review & editing

Hu Zeng: Formal analysis, Writing - review & Editing

Moses Rodriguez: Formal analysis

Cornelia M. Weyand: Formal analysis

John M. Davis III: Conceptualization, Formal analysis, Funding acquisition, Investigation, Project administration, Resources, Supervision, Writing - Review & Editing

Jaeyun Sung: Conceptualization, Formal analysis, Funding acquisition, Investigation, Methodology, Project administration, Supervision, Visualization, Writing - Original Draft, Writing - Review & Editing

## References

[1] J.S. Smolen, D. Aletaha, A. Barton, G.R. Burmester, P. Emery, G.S. Firestein, A. Kavanaugh, I.B. McInnes, D.H. Solomon, V. Strand, K. Yamamoto, Rheumatoid arthritis, Nat. Rev. Dis. Primers. 4 (2018) 18001.

[2] E. Myasoedova, C.S. Crowson, H.M. Kremers, T.M. Therneau, S.E. Gabriel, Is the incidence of rheumatoid arthritis rising?: results from Olmsted County, Minnesota, 1955-2007, Arthritis Rheum. 62 (2010) 1576–1582.

[3] D. Sieghart, A. Platzer, P. Studenic, F. Alasti, M. Grundhuber, S. Swiniarski, T. Horn, H. Haslacher, S. Blüml, J. Smolen, G. Steiner, Determination of autoantibody isotypes increases the sensitivity of serodiagnostics in rheumatoid arthritis, Front. Immunol. 9 (2018). https://doi.org/10.3389/fimmu.2018.00876.

[4] J. Rönnelid, M.C. Wick, J. Lampa, S. Lindblad, B. Nordmark, L. Klareskog, R.F. van Vollenhoven, Longitudinal analysis of citrullinated protein/peptide antibodies (anti-CP) during 5 year follow up in early rheumatoid arthritis: anti-CP status predicts worse disease activity and greater radiological progression, Ann. Rheum. Dis. 64 (2005) 1744–1749.

[5] L.E. Burgers, H.W. van Steenbergen, R.M. Ten Brinck, T.W. Huizinga, A.H. van der Helm-van Mil, Differences in the symptomatic phase preceding ACPA-positive and ACPA-negative RA: a longitudinal study in arthralgia during progression to clinical arthritis, Ann. Rheum. Dis. 76 (2017) 1751–1754.

[6] J. Grosse, E. Allado, C. Roux, A. Pierreisnard, M. Couderc, I. Clerc-Urmes, T. Remen, É. Albuisson, M. De Carvalho-Bittencourt, I. Chary-Valckenaere, D. Loeuille, ACPA-positive versus ACPA-negative rheumatoid arthritis: two distinct erosive disease entities on radiography and ultrasonography, Rheumatol. Int. 40 (2020) 615–624.

[7] S.D. Seegobin, M.H.Y. Ma, C. Dahanayake, A.P. Cope, D.L. Scott, C.M. Lewis, I.C. Scott, ACPA-positive and ACPA-negative rheumatoid arthritis differ in their requirements for combination DMARDs and corticosteroids: secondary analysis of a randomized controlled trial, Arthritis Res. Ther. 16 (2014) R13.

[8] N.A. Daha, R.E.M. Toes, Rheumatoid arthritis: Are ACPA-positive and ACPA-negative RA the same disease?, Nat. Rev. Rheumatol. 7 (2011) 202–203.

[9] L. Padyukov, M. Seielstad, R.T.H. Ong, B. Ding, J. Rönnelid, M. Seddighzadeh, L. Alfredsson, L. Klareskog, Epidemiological Investigation of Rheumatoid Arthritis (EIRA) study group, A genome-wide association study suggests contrasting associations in ACPA-positive versus ACPA-negative rheumatoid arthritis, Ann. Rheum. Dis. 70 (2011) 259–265.

[10] E. Myasoedova, J. Davis, E.L. Matteson, C.S. Crowson, Is the epidemiology of rheumatoid arthritis changing? Results from a population-based incidence study, 1985-2014, Ann. Rheum. Dis. 79 (2020) 440–444.

[11] H. Luan, W. Gu, H. Li, Z. Wang, L. Lu, M. Ke, J. Lu, W. Chen, Z. Lan, Y. Xiao, J. Xu, Y. Zhang, Z. Cai, S. Liu, W. Zhang, Serum metabolomic and lipidomic profiling identifies diagnostic biomarkers for seropositive and seronegative rheumatoid arthritis patients, J. Transl. Med. 19 (2021) 500.

[12] X. Wu, Y. Liu, S. Jin, M. Wang, Y. Jiao, B. Yang, X. Lu, X. Ji, Y. Fei, H. Yang, L. Zhao, H. Chen, Y. Zhang, H. Li, P.E. Lipsky, G.C. Tsokos, F. Bai, X. Zhang, Single-cell sequencing of immune cells from anticitrullinated peptide antibody positive and negative rheumatoid arthritis, Nat. Commun. 12 (2021) 4977.

[13] A. Floudas, M. Canavan, T. McGarry, R. Mullan, S. Nagpal, D.J. Veale, U. Fearon, ACPA status correlates with differential immune profile in patients with rheumatoid arthritis, Cells. 10 (2021) 647.

[14] J. He, Y. Chu, J. Li, Q. Meng, Y. Liu, J. Jin, Y. Wang, J. Wang, B. Huang, L. Shi, X. Shi, J. Tian, Y. Zhufeng, R. Feng, W. Xiao, Y. Gan, J. Guo, C. Shao, Y. Su, F. Hu, X. Sun, J. Yu, Y. Kang, Z. Li, Intestinal butyrate-metabolizing species contribute to autoantibody production and bone erosion in rheumatoid arthritis, Sci. Adv. 8 (2022) eabm1511.

[15] T.B.G. Poulsen, D. Damgaard, M.M. Jørgensen, L. Senolt, J.M. Blackburn, C.H. Nielsen, A. Stensballe, Identification of novel native autoantigens in rheumatoid arthritis, Biomedicines. 8 (2020) 141.

[16] T.B.G. Poulsen, D. Damgaard, M.M. Jørgensen, L. Senolt, J.M. Blackburn, C.H. Nielsen, A. Stensballe, Identification of potential autoantigens in anti-CCP-positive and anti-CCP-negative rheumatoid arthritis using citrulline-specific protein arrays, Sci. Rep. 11 (2021) 17300.

[17] E. Reed, A.K. Hedström, M. Hansson, L. Mathsson-Alm, B. Brynedal, S. Saevarsdottir, M. Cornillet, P.-J. Jakobsson, R. Holmdahl, K. Skriner, G. Serre, L. Alfredsson, J. Rönnelid, K. Lundberg, Presence of autoantibodies in “seronegative” rheumatoid arthritis associates with classical risk factors and high disease activity, Arthritis Res. Ther. 22 (2020) 170.

[18] K. Li, W. Mo, L. Wu, X. Wu, C. Luo, X. Xiao, X. Jia, H. Yang, Y. Fei, H. Chen, F. Zhang, Y. Li, L. Zhao, X. Zhang, Novel autoantibodies identified in ACPA-negative rheumatoid arthritis, Ann. Rheum. Dis. 80 (2021) 739–747.

[19] D. Aletaha, T. Neogi, A.J. Silman, J. Funovits, D.T. Felson, C.O. Bingham 3rd, N.S. Birnbaum, G.R. Burmester, V.P. Bykerk, M.D. Cohen, B. Combe, K.H. Costenbader, M. Dougados, P. Emery, G. Ferraccioli, J.M.W. Hazes, K. Hobbs, T.W.J. Huizinga, A. Kavanaugh, J. Kay, T.K. Kvien, T. Laing, P. Mease, H.A. Ménard, L.W. Moreland, R.L. Naden, T. Pincus, J.S. Smolen, E. Stanislawska-Biernat, D. Symmons, P.P. Tak, K.S. Upchurch, J. Vencovský, F. Wolfe, G. Hawker, 2010 Rheumatoid arthritis classification criteria: an American College of Rheumatology/European League Against Rheumatism collaborative initiative, Arthritis Rheum. 62 (2010) 2569–2581.

[20] E. von Elm, D.G. Altman, M. Egger, S.J. Pocock, P.C. Gøtzsche, J.P. Vandenbroucke, STROBE Initiative, Strengthening the Reporting of Observational Studies in Epidemiology (STROBE) statement: guidelines for reporting observational studies, BMJ. 335 (2007) 806–808.

[21] D. Aletaha, J. Smolen, The Simplified Disease Activity Index (SDAI) and the Clinical Disease Activity Index (CDAI): a review of their usefulness and validity in rheumatoid arthritis, Clin. Exp. Rheumatol. 23 (2005) S100–8.

[22] N. Cliff, Ordinal methods for behavioral data analysis, Psychology Press, London, England, 2016.

[23] D.W. Huang, B.T. Sherman, R.A. Lempicki, Systematic and integrative analysis of large gene lists using DAVID bioinformatics resources, Nat. Protoc. 4 (2009) 44–57.

[24] T.W. Kragstrup, K. Otkjaer, C. Holm, A. Jørgensen, M. Hokland, L. Iversen, B. Deleuran, The expression of IL-20 and IL-24 and their shared receptors are increased in rheumatoid arthritis and spondyloarthropathy, Cytokine. 41 (2008) 16–23.

[25] E.E. Davenport, T. Amariuta, M. Gutierrez-Arcelus, K. Slowikowski, H.-J. Westra, Y. Luo, C. Shen, D.A. Rao, Y. Zhang, S. Pearson, D. von Schack, J.S. Beebe, N. Bing, S. John, M.S. Vincent, B. Zhang, S. Raychaudhuri, Discovering in vivo cytokine-eQTL interactions from a lupus clinical trial, Genome Biol. 19 (2018) 168.

[26] J. Wang, L. Fang, L. Ye, S. Ma, H. Huang, X. Lan, J. Ma, miR-137 targets the inhibition of TCF4 to reverse the progression of osteoarthritis through the AMPK/NF-κB signaling pathway, Biosci. Rep. 40 (2020). https://doi.org/10.1042/BSR20200466.

[27] K.S. Ravichandran, U. Lorenz, Engulfment of apoptotic cells: signals for a good meal, Nat. Rev. Immunol. 7 (2007) 964–974.

[28] A. Lleo, C. Selmi, P. Invernizzi, M. Podda, M.E. Gershwin, The consequences of apoptosis in autoimmunity, J. Autoimmun. 31 (2008) 257–262.

[29] M. Kalaaji, E. Mortensen, L. Jørgensen, R. Olsen, O.P. Rekvig, Nephritogenic lupus antibodies recognize glomerular basement membrane-associated chromatin fragments released from apoptotic intraglomerular cells, Am. J. Pathol. 168 (2006) 1779–1792.

[30] O. Cabral-Marques, A. Marques, L.M. Giil, R. De Vito, J. Rademacher, J. Günther, T. Lange, J.Y. Humrich, S. Klapa, S. Schinke, L.F. Schimke, G. Marschner, S. Pitann, S. Adler, R. Dechend, D.N. Müller, I. Braicu, J. Sehouli, K. Schulze-Forster, T. Trippel, C. Scheibenbogen, A. Staff, P.R. Mertens, M. Löbel, J. Mastroianni, C. Plattfaut, F. Gieseler, D. Dragun, B.E. Engelhardt, M.J. Fernandez-Cabezudo, H.D. Ochs, B.K. Al-Ramadi, P. Lamprecht, A. Mueller, H. Heidecke, G. Riemekasten, GPCR-specific autoantibody signatures are associated with physiological and pathological immune homeostasis, Nat. Commun. 9 (2018) 5224.

[31] S. Nagata, Apoptosis mediated by the Fas system, Prog. Mol. Subcell. Biol. 16 (1996) 87–103.

[32] C.T. Chou, J.S. Yang, M.R. Lee, Apoptosis in rheumatoid arthritis--expression of Fas, Fas-L, p53, and Bcl-2 in rheumatoid synovial tissues, J. Pathol. 193 (2001) 110–116.

[33] L. Peng, N. Zhu, J. Mao, L. Huang, Y. Yang, Z. Zhou, L. Wang, B. Wu, Expression levels of CXCR4 and CXCL12 in patients with rheumatoid arthritis and its correlation with disease activity, Exp. Ther. Med. 20 (2020) 1925–1934.

[34] A. Mak, N.Y. Kow, N.H. Ismail, N.D. Anuar, N.H. Rutt, J. Cho, N.S.B.M. Rosli, B. Dharmahidari, Detection of putative autoantibodies in systemic lupus erythematous using a novel native-conformation protein microarray platform, Lupus. 29 (2020) 1948–1954.

[35] S. Vordenbäumen, P. Böhmer, R. Brinks, R. Fischer-Betz, J. Richter, E. Bleck, P. Rengers, H. Göhler, H.-D. Zucht, P. Budde, P. Schulz-Knappe, M. Schneider, High diagnostic accuracy of histone H4-IgG autoantibodies in systemic lupus erythematosus, Rheumatology (Oxford). 57 (2018) 533–537.

[36] O. Cabral-Marques, G. Halpert, L.F. Schimke, Y. Ostrinski, A. Vojdani, G.C. Baiocchi, P.P. Freire, I.S. Filgueiras, I. Zyskind, M.T. Lattin, F. Tran, S. Schreiber, A.H.C. Marques, D.R. Plaça, D.L.M. Fonseca, J.Y. Humrich, A. Müller, L.M. Giil, H. Graßhoff, A. Schumann, A. Hackel, J. Junker, C. Meyer, H.D. Ochs, Y.B. Lavi, C. Scheibenbogen, R. Dechend, I. Jurisica, K. Schulze-Forster, J.I. Silverberg, H. Amital, J. Zimmerman, H. Heidecke, A.Z. Rosenberg, G. Riemekasten, Y. Shoenfeld, Autoantibodies targeting GPCRs and RAS-related molecules associate with COVID-19 severity, Nat. Commun. 13 (2022) 1220.

[37] E.C. Farlow, K. Patel, S. Basu, B.-S. Lee, A.W. Kim, J.S. Coon, L.P. Faber, P. Bonomi, M.J. Liptay, J.A. Borgia, Development of a multiplexed tumor-associated autoantibody-based blood test for the detection of non-small cell lung cancer, Clin. Cancer Res. 16 (2010) 3452–3462.

[38] F. Le Naour, F. Brichory, D.E. Misek, C. Bréchot, S.M. Hanash, L. Beretta, A distinct repertoire of autoantibodies in hepatocellular carcinoma identified by proteomic analysis, Mol. Cell. Proteomics. 1 (2002) 197–203.

[39] S. Gnjatic, E. Ritter, M.W. Büchler, N.A. Giese, B. Brors, C. Frei, A. Murray, N. Halama, I. Zörnig, Y.-T. Chen, C. Andrews, G. Ritter, L.J. Old, K. Odunsi, D. Jäger, Seromic profiling of ovarian and pancreatic cancer, Proc. Natl. Acad. Sci. U. S. A. 107 (2010) 5088–5093.

[40] W. Katchamart, A. Koolvisoot, E. Aromdee, P. Chiowchanwesawakit, C. Muengchan, Associations of rheumatoid factor and anti-citrullinated peptide antibody with disease progression and treatment outcomes in patients with rheumatoid arthritis, Rheumatol. Int. 35 (2015) 1693–1699.

[41] I.B. McInnes, G. Schett, The pathogenesis of rheumatoid arthritis, N. Engl. J. Med. 365 (2011) 2205–2219.

[42] Y. Shoenfeld, E. Toubi, Protective autoantibodies: role in homeostasis, clinical importance, and therapeutic potential, Arthritis Rheum. 52 (2005) 2599–2606.

[43] H.U. Lutz, C.J. Binder, S. Kaveri, Naturally occurring auto-antibodies in homeostasis and disease, Trends Immunol. 30 (2009) 43–51.

[44] B. Wootla, S. Lacroix-Desmazes, A.E. Warrington, A.J. Bieber, S.V. Kaveri, M. Rodriguez, Autoantibodies with enzymatic properties in human autoimmune diseases, J. Autoimmun. 37 (2011) 144–150.

[45] C. Grönwall, G.J. Silverman, Natural IgM: beneficial autoantibodies for the control of inflammatory and autoimmune disease, J. Clin. Immunol. 34 Suppl 1 (2014) S12–21.

[46] G.J. Silverman, J. Vas, C. Grönwall, Protective autoantibodies in the rheumatic diseases: lessons for therapy, Nat. Rev. Rheumatol. 9 (2013) 291–300.

[47] M. Neiman, C. Hellström, D. Just, C. Mattsson, L. Fagerberg, I. Schuppe-Koistinen, A. Gummesson, G. Bergström, O. Kallioniemi, A. Achour, R. Sallinen, M. Uhlén, P. Nilsson, Individual and stable autoantibody repertoires in healthy individuals, Autoimmunity. 52 (2019) 1–11.

[48] M. Shome, Y. Chung, R. Chavan, J.G. Park, J. Qiu, J. LaBaer, Serum autoantibodyome reveals that healthy individuals share common autoantibodies, Cell Rep. 39 (2022) 110873.

[49] Y. Xu, Q. Wu, Prevalence trend and disparities in rheumatoid arthritis among US adults, 2005-2018, J. Clin. Med. 10 (2021) 3289.

[50] G. Barturen, L. Beretta, R. Cervera, R. Van Vollenhoven, M.E. Alarcón-Riquelme, Moving towards a molecular taxonomy of autoimmune rheumatic diseases, Nat. Rev. Rheumatol. 14 (2018) 180.

[51] J. Sung, Y. Wang, S. Chandrasekaran, D.M. Witten, N.D. Price, Molecular signatures from omics data: from chaos to consensus, Biotechnol. J. 7 (2012) 946–957.

[52] V.K. Gupta, K.Y. Cunningham, B. Hur, U. Bakshi, H. Huang, K.J. Warrington, V. Taneja, E. Myasoedova, J.M. Davis 3rd, J. Sung, Gut microbial determinants of clinically important improvement in patients with rheumatoid arthritis, Genome Med. 13 (2021) 149.

[53] B. Hur, V.K. Gupta, H. Huang, K.A. Wright, K.J. Warrington, V. Taneja, J.M. Davis 3rd, J. Sung, Plasma metabolomic profiling in patients with rheumatoid arthritis identifies biochemical features predictive of quantitative disease activity, Arthritis Res. Ther. 23 (2021) 164.

[54] V.K. Gupta, M. Kim, U. Bakshi, K.Y. Cunningham, J.M. Davis 3rd, K.N. Lazaridis, H. Nelson, N. Chia, J. Sung, A predictive index for health status using species-level gut microbiome profiling, Nat. Commun. 11 (2020) 4635.

[55] C.M. Coffey, C.S. Crowson, E. Myasoedova, E.L. Matteson, J.M. Davis 3rd, Evidence of diagnostic and treatment delay in seronegative rheumatoid arthritis: Missing the window of opportunity, Mayo Clin. Proc. 94 (2019) 2241–2248.

[56] E.A. Littlejohn, S.U. Monrad, Early diagnosis and treatment of rheumatoid arthritis, Prim. Care. 45 (2018) 237–255.

