## Supplementary Information for "Patients with ACPA-positive and ACPA-negative Rheumatoid Arthritis Show Different Serological Autoantibody Repertoires and Autoantibody Associations with Disease Activity"

for

Kevin Y. Cunningham *et al.*

#### Data handling and pre-processing

For each slide, proteins and control probes are in quadruplicates, with 4 arrays on each slide.

**Step 1:** Calculate the net intensities for each spot by subtracting the background signal intensities from the foreground signal intensities of each spot. For each spot, the background signal intensity is calculated using a circular region that is centered on the spot. **Supplementary Fig. 1** is a visual representation of how this calculation was carried out (*Molecular Devices*).

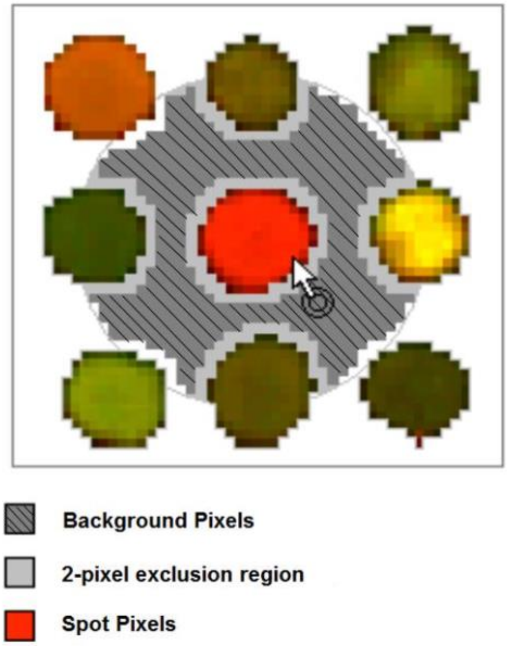


**Supplementary Figure 1.** Visual representation of the local background calculation methods. The red spot in the middle is the spot of interest.

**Step 2:** Remove replica spots with an RFU $\leq 0$.

**Step 3:** Zero net intensities if only 1 replica spot remaining.

**Step 4:** Calculate the percentage of coefficient of variant (CV%) to determine the variations between the replica spots on each slide.

$$\text{CV\%} = \frac{\text{S.D.}}{\text{Mean}} \times 100\%$$

Flag a set of replica spots with only 2 or less replica/s remaining and CV% > 20% as "High CV". The mean RFU of these replica spots (proteins) will be excluded from the downstream analysis.

For the proteins and controls with a CV% > 20% and with 3 or more replica spots remaining, the replica spots that resulted in this high CV% value were filtered out. This was done by first calculating the standard deviation between the median value of the net intensities and the individual net intensities for each set of the replica spots.

The spot with the largest standard deviation was removed. CV% values were re-calculated and the process repeated.

**Step 5:** Calculate the mean of the net intensities for the remaining replica spots.

**Step 6:** Inspect the signal intensities of the two positive controls: IgG and Cy3-BSA.

**Step 7:** Composite normalization of data using both quantile-based and total intensity-based modules (Duarte, J. *et al.*, 2013). This method assumes that the different samples share a common distribution of their control probes while taking into account the potential existence of flagged spots within them. The Immunome array uses Cy3-labeled biotinylated BSA (Cy3-BSA) replicates as the positive control spots across slides. It is considered a housekeeping probe for normalization of signal intensities for any given study.

The quantile module adopts the algorithm in Bolstad *et al.*, 2003. This enables the detection and handling of outliers or flagged spots in any of the Cy3-BSA control probes. A total intensity-based module was then implemented to obtain a scaling factor for each sample. This assumes that post-normalization, the positive controls should have a common total intensity value across all samples (Causton H.C. *et al.*, 2004). This method aims to normalize the protein array data from variations in their measurements while preserving the targeted biological activity across samples.

**Quantile-based normalization of all Cy3-BSA across all samples**

(i = spot number and j = sample number)

1. Load all Cy3-BSA across all samples, j, into an $i \times j$ matrix X
2. Sort spot intensities in each column j of X to get $X_{sort}$
3. Take the mean across each row i of $X_{sort}$ to get $< X_i >$

**Intensity-based normalization**

1. Calculate the sum of the mean across each row i, $\sum < X_i >_i$
2. For each sample, k, calculate the sum of all Cy3-BSA controls, $\sum X_k$
3. For each sample, k,

$$\text{Scaling factor(k)} = \frac{\sum <Xi>}{\sum Xk}$$

### References

1. Bolstad, B. M., Irizarry, R. A., Astrand, M. & Speed, T. P. (2003). A Comparison of Normalization Methods for High Density Oligonucleotide Array Data Based on Bias and Variance. Bioinformatics. 19(2):185-193.
2. Causton, H.C., Quackenbush, J. & Brazma, A. (2004). Microarray gene expression data analysis: a beginners' guide. 1st ed. Malden: Blackwell Publishing.
3. Duarte, J. et al. (2013). Protein function microarrays: design, use, and bioinformatic analysis in cancer biomarker discovery and quantitation. Bioinformatics of Human Proteomics, Springer Netherlands. 39-74.
4. Molecular Devices, GenePix Pro 7 Software User Guide, 0112-0178 Rev B (June 2010) Figure 1-1 p25.

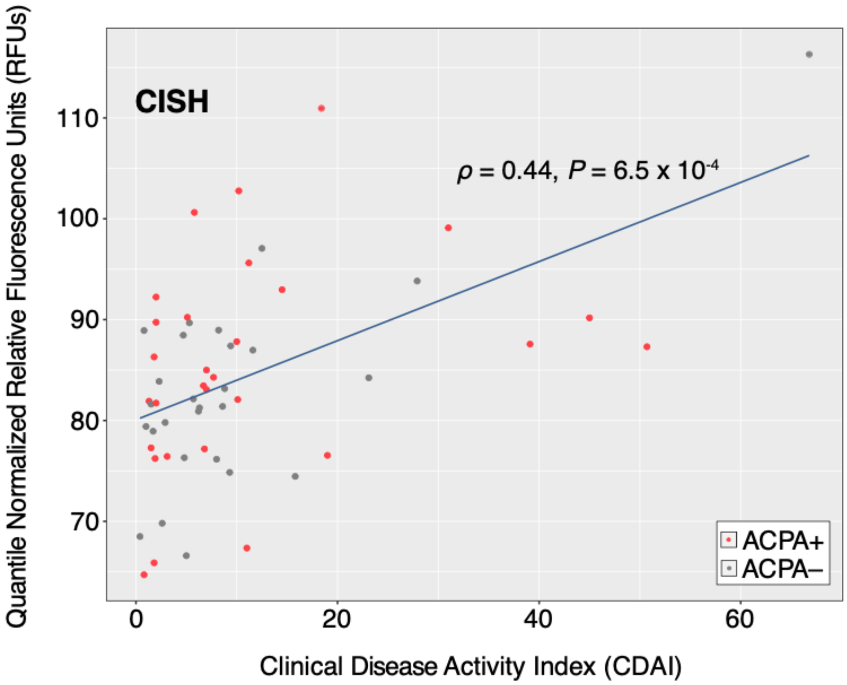


**Supplementary Figure 2. Autoantibodies targeting CISH proteins are positively correlated with Clinical Disease Activity Index (CDAI) (|Spearman's $\rho$| > 0.44 and $P$ = 6.5 × $10^{-4}$) when considering both RA subgroups simultaneously, but not in either ACPA+ RA ($\rho$ = 0.46, $P$ = 0.0108) or ACPA– RA ($\rho$ = 0.39, $P$ = 0.04) separately.** The blue line in the scatterplots represents the linear regression fit of Y~X, wherein X and Y correspond to CDAI and autoantibody abundance, respectively.
